# Taking a participatory research approach within workplace health promotion research to improve physical activity levels in office-based workers: a scoping review protocol

**DOI:** 10.1101/2021.09.01.21262961

**Authors:** Aidan Buffey, Brian P Carson, Alan Donnelly, Jon Salsberg

## Abstract

**Introduction:** Physical activity (PA) workplace health promotion (WHP) interventions have traditionally utilised a top-down research approach at an individual level where participants are considered as passive subjects. Whereas participatory research involves the participants and relevant stakeholders within the research process utilising a bottom-up approach which focuses on the health priorities of the participants and allows the integration of the researcher’s expertise and the end-users lived experiences, which has been shown to aid in the acceptability and relevance of the research.

This protocol describes a scoping review which will explore, identify and map participatory research techniques and their impact when utilised in office based WHP interventions designed to improve PA levels and/or decrease sitting time. Providing an overview of key characteristics of WHP interventions which took a participatory research approach.

**Methods and analysis:** This scoping review will follow the guidelines and framework from the PRISMA-ScR. Articles will be retrieved via five databases: Web of Science, PubMED, Scopus, Google Scholar and OpenGrey. A search strategy was piloted, and relevant review articles search strategies were explored, to identify appropriate key words and MeSH terms. Two independent reviewers will screen retrieved articles based on our inclusion and exclusion criteria by title and abstract first, followed by the full text. Any discrepancies will be discussed until a consensus is reached. Data will be extracted, charted and summarised via a narrative synthesis and qualitative analyses.

**Ethics and dissemination:** Ethical approval was not required or obtained for this scoping review. The completed scoping review findings will be disseminated in a peer-reviewed journal which has a research scope that encompasses participatory research and health promotion. The findings will be presented at appropriate academic conferences and to project partners to inform the design of a WHP intervention.

**Strengths and limitations of this study:**

- The proposed scoping review will explore and map the current participatory research techniques and approaches taken when completing an office-based workplace health promotion intervention designed to increase physical activity.
- The scoping review by nature and the search strategy proposed will allow for a wide breadth of literature to be explored.
- There is no critical appraisal or quality assessment of the included studies which is typical of a systematic review but not of a scoping review or the aim of this review article.
- This scoping review has a narrow focus on physical activity and may limit/exclude articles aiming to improve psychological wellbeing or work performance.

## Introduction

Within occupational health research, prolonged occupational sitting is a topic of increased debate and growing research.(1) Changes in the workplace environment have been associated with significant reductions in the demand of physical activity (PA) and the increased use of computers has shown an increased prevalence of prolonged sitting in many workplaces, especially office-based workplaces.(2-3) Sitting is a sedentary behaviour (SB) which is a term used to classify low levels of energy expenditure when sitting or in a reclined posture.(3) The workplace environment and organisational culture can often facilitate and promote prolonged SB.(2, 4) Two previous studies objectively measured sedentary time, using accelerometers, in office workers.(5-6) They found office workers were sedentary for a mean of 75.8% (95% CI: 74.5, 77.1)(6) and 81.8% (438.8 ± 51.5 minutes)(5) during working hours.

Previous workplace health promotion (WHP) interventions have taken a traditional top-down research driven approach where participants or communities are considered as passive subjects.(7-8) Whereas participatory research (PR) is an approach to research that involves the target population and relevant stake holders within the research process to promote and establish a sense of ownership which helps to increase the relevance of the research.(7) This sense of ownership is seen in health promotion (HP) research which at its philosophical core focuses on empowerment and community participation.(9) An early description of HP by the World Health Organisation establishes HP as a method of empowering individuals and communities to take control over their health and its determinants.(10)

Many WHP interventions have been conducted and targeted different aspects of either the workplace environment, workhours schedule, transport to and from work or targeted behavioural changes to increase PA and/or decrease sitting time. Some examples are (a) taking the stairs, in place of an elevator, (b) encourage achieving the recommended moderate-to-vigorous PA guidelines, (c) active e-mails and or walking meetings, (d) active transport (walking or cycling to and from work), (e) height adjustable desks, treadmill desks, cycling desks, (f) pedometers/accelerometers that measure PA and (g) breaking sitting time.(1, 3, 5, 11) These interventions however are typically administered at an individual level with a traditional top-down research approach. Previous WHP interventions that did not implement a participative approach to the intervention when targeting behaviour change have been shown to be weaker in design.(12) Whilst workplace interventions that employed a participatory approach accounting for factors such as individual, interpersonal, organisational, workplace community, company policy and workplace environment and the interaction between these factors are considered multilevel interventions.(1) It has been proposed that multilevel interventions are typically more effective than individual level interventions when aiming to increase moderate to vigorous PA.(1) Multilevel interventions conducted with a PR approach have been shown to be beneficial in numerous ways. As the inclusion of the target population compliments the research process and offers real-world viewpoints, practical solutions and can aid in the translation of discovered knowledge into practice; it is also suggested that the PR process improves participation and enthusiasm towards an intervention.(13-15)

Creating a PA WHP intervention which is sustainable after the completion of the study should be a research priority and maintaining participants adherence during the intervention can be difficult, with high rates of attrition shown in previous WHP studies.(11) For example, participants who are highly sedentary prior to an intervention are likely to return to their previous sedentary behaviour, due to increasing work pressures.(2) Therefore, studies utilising a PR approach which fosters a motivational component to the study for the participant may be beneficial.(2) PR and the HP movement both begin with the health priorities of the individuals or community with a bottom-up approach, with the philosophy that those who are affected by the research should have a say in what and how research is conducted.(15-17) This inclusivity with the participants, with the aim of collaboration, education and community action promotes active involvement within the research process.(7, 15-16)

PR allows the integration of the researchers’ expertise and the target populations lived experiences which is a significant strength of the PR approach to research and has been shown to aid the acceptability of the research when implemented if conducted well.(13) The PR approach has been shown to be an influential tool at multiple levels of the research process through the inclusion of community members as collaborators.(15) This strength of PR research has led to an increase in academics utilising the PR methodology as HP needs to be implemented successfully for an improvement in health to be observed.(8, 18)

### Rationale

Previous research investigating work site health has taken traditional research approaches such as in-person interventions, printed materials and information talks.(19) Malik et al.(11) completed a systematic review of WHP interventions designed to promote PA and the impact they had on participant’s PA levels. They found evidence to suggest WHP interventions can be effective and showed positive outcomes for some of the included studies, however the overall results were inconclusive and called for more research into the elements of WHP interventions that are likely to increase efficacy and adoption within the workplace.(11)

Taking a PR approach can take various forms with varying methodologies and can impact the effectiveness of the WHP intervention. When a participatory approach is not taken it has been shown to lead to inappropriateness of an intervention approach/concept or format.(20) Previous literature has examined the benefits and effectiveness of WHP interventions. However, to the authors knowledge, the use of PR within WHP intervention has not been synthesised. Therefore, the authors aimed to examine how PR is being incorporated within WHP research, to identify the current available evidence, key methods, and the scope of reported impacts of PR. Providing an overview and identifying key characteristics of the current research that has utilised PR within the WHP intervention. A scoping review was therefore identified as the most appropriate methodological approach to investigate the use of PR within WHP interventions and to map and explore all available evidence and identify and analyse any knowledge gaps.(21)

### Research Aims

This scoping review aims to identify current research practices when taking a PR approach to WHP interventions aimed to increase PA and/or reduce sitting time and the impact the PR approach has on the research. Due to the broad nature of the scoping review, we developed key research objectives that we believe address the overall aim of the review.

### Objectives

The overall objectives of the scoping review relate to identifying and mapping previous literature to provide a base of evidence for researchers who plan to take a PR approach in future WHP interventions. Our objectives are:

1. Identify and map previous literature where office-based adults have been involved in PR studies and how their involvement shaped the design of the WHP intervention.
2. Identify and discuss the methods implemented in the WHP studies that took a PR approach.
3. Discuss the evaluation and outcomes measured in the WHP articles included in the scoping review that took a PR approach.

## Method

### Protocol

The scoping review will follow the guidelines and framework from the Preferred Reporting Items for Systematic reviews and Meta-Analyses extension for Scoping Reviews (PRISMA-ScR).(22) The PRISMA-ScR consists of a 22-item checklist (Found: http://www.prisma-statement.org/documents/PRISMA-ScR-Fillable-Checklist_11Sept2019.pdf).

### Information sources

A systematic search spanning five electronic databases, which includes Web of Science, PubMED, Scopus, Google Scholar and OpenGrey will be completed between the years of 1995-2021. Articles will be screened for eligibility relating to our inclusion and exclusion criteria (See Table 1). After the removal of duplicated articles, two reviewers will first screen the title and abstract of the retrieved items for eligibility. Selection will then be confirmed by two reviewers after screening the full text, with any disagreement being resolved between reviewers, with arbitration where needed by a third reviewer, until a consensus is reached. Grey literature will be screened from Google Scholar and OpenGrey in the same process as the articles retrieved from Web of Science, PubMED and Scopus. The references of the included studies will be screened, and we will look to include any relevant grey literature. The PRISMA flow diagram template will be published alongside the scoping review to illustrate the search strategy screening process, providing the number of sources screened, with reasons for exclusion and the final number of included studies.(22)

**Table 1.** Eligibility criteria. Illustrates the eligibility criteria, with the inclusion and exclusion criteria and rationale statements.

| Characteristics | Inclusion | Exclusion | Rationale |
| --- | --- | --- | --- |
| **Population** | Humans.<br>Working adults.<br>Office environment. | Animal studies.<br>Non-working adults.<br>Children, teenagers, and retired adults.<br>Non-office or home-based workers. | 1. The focus of this scoping review is to investigate PR in WHP studies in office-based participants/workplaces.<br>2. Children, teenagers and retired adults would not fit our eligibility criteria of ‘working adults’.<br>3. Non-office workers may have different ‘health’ needs relating to the working environment. |
| **Language** | English language | All other languages. | 1. The reviewers only speak English. |
| **Years considered** | 1995-2021 | Years outside this time period. | 1. A wide time period to capture all relevant WHP research.<br>2. PR became more prominent in HP studies in 1995 and therefore implemented into research practices following this year. |
| **Study focus** | 1. Articles investigating workplace health promotion in office-based workplaces that implemented participatory health research techniques.<br>2. Includes a physical activity aspect to the intervention study, for example increasing physical activity or decreasing sitting time. | Studies must be conducted within the workplace with the aim of improving health.<br>Not based in the community or home.<br>The health promotion intervention was also measuring/targeting psychological or work-performance improvements. | 1. The focus of the overall research question of this scoping review, is specific to PR in WHP research in office-based workplaces.<br>2. Other work-based environments may carry different health associated risks, priorities or safety concerns which would not be comparable to that of an office-based environment.<br>3. Including outcome measures will allow for an evaluation of the included studies and a discussion on the effectiveness of taking a PR approach in those WHP studies. |
| **Publication status** | Published peer-reviewed journal articles and relevant grey literature. Relevant grey literature is defined within this scoping review as: theses/dissertations, conference papers, research/government reports, ongoing research, editorials, and textbooks. | Any other literature that is not listed in the inclusion criteria, such as websites. | 1. The aim of this scoping review is to capture a wide breadth of literature so including grey literature insures a more complete search and minimises publication bias. |

### Search strategy and eligibility criteria

To best capture the breadth of literature that we were hoping to retrieve that fit our eligibility criteria (See Table 1), we piloted preliminary searches and referred to previous review articles search terms in the research area of WHP interventions.(11) After performing preliminary searches of the Web of Science database and identifying key words from article titles and abstracts, we identified the MeSH terms of these keywords using PubMED. These keywords and MeSH terms were then used across all included databases and adapted where needed across the databases. A complete search strategy illustrating the search strategy used for Web of Science is included in Supplementary File 1.

### Data Charting

Data from the retained studies will be charted independently by one reviewer with a sample of the included studies being duplicated by a second reviewer independently to confirm the data charting process. Any discrepancies will be discussed until a consensus is researched, arbitrated as needed by a third reviewer, and the data charting process will be confirmed. Data will be extracted from the included studies and charted into a Microsoft Excel sheet [Microsoft Excel, 2011] table. The data extracted will be charted into the Microsoft Excel sheet following the headings shown below and filled in with information answering the associated questions (See Table 2).

**Table 2.** Data charting. Table displaying the data charting headings and associated questions used to retrieve information from the included articles and extracted.

| **Table 2.** | **Data charting** |
| --- | --- |
| **Charted Data** | **Associated Questions** |
| **First author** | What is the name of the first author? |
| **Title of journal article** | What is the title of the published article? |
| **Year of publication** | What year was the article published? |
| **Origin** | Where was the study conducted? |
| **Population** | What are the descriptive characteristics of the studies participant sample group? |
| **Study design** | What methodological design did the researchers utilise? |
| **Study purpose and aims** | What was the purpose of the study?<br>What were the aims/objectives of the study? |
| **Study procedures** | What was the length of the study?<br>When and how were measures taken (baseline, outcomes, follow up)? |
| **Participatory research techniques** | What participatory research techniques are mentioned, detailed, and implemented within the methodology of the study?<br>How and where within the study were the participatory research techniques implemented? |
| **Oversight** | Was there any oversight to the intervention, specifically a participatory participant group separate or built into the researcher team? |
| **Intervention focus** | Was there an intervention? What did the intervention focus on/measure?<br>What was the physical activity intervention? Methods and details. |
| **Data collection** | How was data collected? Note down recorded findings. |
| **Study outcomes** | What were the study outcomes? Note down the reported study outcomes. |
| **Data analysis** | How was data analysed? Was a PR approach taken in regard to data analysis or data checking? |
| **Evaluation of PR techniques** | Did the WHP intervention evaluate any of the PR techniques implemented? If so, how? Note down their self-evaluation.<br>Following Jagosh et al.(23), self-reported PR was assessed by asking of the article; ‘Does the full-text paper indicate that participation occurred in the following three areas: (A) Partners were involved in identifying or setting the research questions? (B) Partners were involved in setting the methodology or collecting data or analysing the data? (C) Partners were involved in uptake or dissemination of the research findings (this requirement was loosely applied because publication often predates uptake)? |
| **Notes** | Any information that may be useful that does not fit into above question asked of the retrieved studies? |

### Synthesis of Results

Following data extraction and charting we will provide a narrative synthesis of the included studies, descriptively summarising the data that has been charted. We will not critically appraise the data; we will look to aggregate the findings of the included studies allowing us to summarise and identify recurring themes. These themes will be reported qualitatively and displayed in a way which answers the proposed research question and objectives.

Where data has been extracted relating to changes in PA, whether negative, positive or neutral, we will present these findings descriptively. We will discuss the methods and characteristics of the studies related to the change in PA.

This narrative synthesis of results will therefore map the existing literature which has taken a PR approach when conducting a WHP intervention and identify the available evidence and impact of taking a PR approach. This scoping review will inform future WHP interventions and provide a base of current evidence for the methods and usages of PR within WHP studies.

### Research implications for future research, practice and policy

By understanding how PR has been implemented and evaluated with WHP interventions, we expect the findings from this scoping review will inform future research questions and indicate the key methods when implementing PR within WHP interventions and the scope of reported impacts of PR.

### Consultation with knowledge users

This scoping review will be presented during the planning stage of a clustered randomised WHP study. As part of the formation of the study, this scoping review will be used to inform and guide stakeholders of the project when planning which and how the PR approach will be taken.

## Supporting information

Supplementary File - Web of Science - Search Stratergy

## Data Availability

No data is currently available with this scoping review protocol.

## Ethics and Dissemination

### Ethics

Ethical approval was/is not required or obtained for this scoping review protocol or the scoping review.

### Dissemination

We plan to disseminate the findings of the completed scoping review through publication, in a peer reviewed journal that incorporates participatory research and health promotion within the scope of the journal as well as presenting the findings at appropriate academic conferences. The findings of the completed scoping review will be presented to project partners and stakeholders during the planning of a WHP intervention.

### Authors’ contributions

All authors have made substantial intellectual contributions to the development of this protocol. AB and JS conceptualised the review approach and AB drafted the manuscript. BC, AD and JS contributed to the conceptualisation, writing and editing of the protocol.

### Funding

This research was supported by the School of Medicine, University of Limerick and a University of Limerick, Health Research Institute Studentship.

## References

1. Plotnikoff R, and Karunamuni N. Reducing sitting time: the new workplace health priority. Arch Environ Occup Health 2012;67:3:125–27 doi:10.1080/19338244.2012.697407 [published Online First: 31 July 2012].

2. Brown DK, Barton JL, Pretty J, et al. Walks4Work: Assessing the role of the natural environment in a workplace physical activity intervention. Scand J Work Environ Health 2014;40:4:390–99 doi:10.5271/sjweh.3421 [published Online First: 13 March 2013].

3. Owen N, Healy GN, Matthews CE, et al. Too much sitting: the population-health science of sedentary behavior. Exerc Sport Sci Rev 2010;38:3:105–13 doi:10.1097/JES.0b013e3181e373a2 [published Online First: 25 July 2012].

4. Marshall AL. Challenges and opportunities for promoting physical activity in the workplace. J Sci Med Sport 2004;7:1:60–6 doi:10.1016/S1440-2440(04)80279-2 [published Online First: 22 February 2006].

5. Parry S, and Straker L. The contribution of office work to sedentary behaviour associated risk. BMC Public Health 2013;13:1:1–10 doi:10.1186/1471-2458-13-296 [published Online First: 04 April 2013]

6. Thorp AA, Healy GN, Winkler E, et al. Prolonged sedentary time and physical activity in workplace and non-work contexts: a cross-sectional study of office, customer service and call centre employees. Int J Behav Nutr Phys Act 2012;9:1:1–9 doi: 10.1186/1479-5868-9-128 [published Online First: 26 October 2012].

7. Macaulay AC, Commanda LE, Freeman WL, et al. Participatory research maximises community and lay involvement. BMJ 1999;319:7212:774–78 doi: 10.1136/bmj.319.7212.774 [published Online First: 18 September 1999].

8. Potvin L, Cargo M, McComber AM, et al. Implementing participatory intervention and research in communities: lessons from the Kahnawake Schools Diabetes Prevention Project in Canada. Social Science & Medicine 2003;56:6:1295–305 doi: 10.1016/S0277-9536(02)00129-6 [published Online First: 14 May 2002].

9. Robertson A, and Minkler M. New health promotion movement: a critical examination. Health Educ Q 1994;21:3:295–312 doi: 10.1177/109019819402100303 [published Online First: 1 October 1994].

10. World Health Organisation. Discussion document on the concept and principles of health promotion. July 9-13, 1984. Copenhagen: European Office of the World Health Organization.

11. Malik SH, Blake H, and Suggs LS. A systematic review of workplace health promotion interventions for increasing physical activity. Br J Health Psychol 2014;19:1:149–80 doi: 10.1111/bjhp.12052 [published Online First: 4 July 2013].

12. Parry S, Straker L, Gilson ND, et al. Participatory workplace interventions can reduce sedentary time for office workers—a randomised controlled trial. PLoS One 2013;8:11:e78957 doi: 10.1371/journal.pone.0078957 [published Online First: 12 November 2013].

13. Cargo M, and Mercer SL. The value and challenges of participatory research: strengthening its practice. Annu Rev Public Health 2008;29:325–50 doi: 10.1146/annurev.publhealth.29.091307.083824 [published Online First: 3 January 2008].

14. Jagosh J, Macaulay AC, Pluye P, et al. Uncovering the benefits of participatory research: implications of a realist review for health research and practice. Milbank Q 2012;90:2:311–46 doi: 10.1111/j.1468-0009.2012.00665.x [published Online First: 18 June 2012].

15. De Las Nueces D, Hacker K, DiGirolamo A, et al. A systematic review of community-based participatory research to enhance clinical trials in racial and ethnic minority groups. Health Serv Res 2012;47:3pt2:1363–86 doi: 10.1111/j.1475-6773.2012.01386.x [first Published Online: 21 February 2012].

16. Shimmin C, Wittmeier KD, Lavoie JG, et al. Moving towards a more inclusive patient and public involvement in health research paradigm: the incorporation of a trauma-informed intersectional analysis. BMC Health Serv Res 2017;17:1:1–10 doi: 10.1186/s12913-017-2463-1 [first Published Online: 7 August 2017].

17. Wallerstein N, and Duran B. Community-based participatory research contributions to intervention research: the intersection of science and practice to improve health equity. A J Public Health 2010;100:S1:S40–6 doi: 10.2105/AJPH.2009.184036 [first Published Online: 20 September 2011].

18. Green LW, Kreuter MW. Health promotion planning: An educational and ecological approach, 4th Edition. New York, McGraw-Hill; 2005.

19. Cook R, Billings D, Hersch R, et al. A field test of a web-based workplace health promotion program to improve dietary practices, reduce stress, and increase physical activity: randomized controlled trial. J Med Internet Res 2007;9:2:e17 doi: 10.2196/jmir.9.2.e17 [first Published Online: 19 June 2007].

20. Rojatz D, Merchant A, and Nitsch M. Factors influencing workplace health promotion intervention: a qualitative systematic review. Health Promot Int, 2017;32:5:831–39 doi: 10.1093/heapro/daw015 [first Published Online: 22 March 2016].

21. Munn Z, Peters MD, Stern C, et al. Systematic review or scoping review? Guidance for authors when choosing between a systematic or scoping review approach. BMC Med Res Methodol 2018;18:1:1–7 doi: 10.1186/s12874-018-0611-x [first Published Online: 19 November 2018].

22. Tricco AC, Lillie E, Zarin W, et al. PRISMA extension for scoping reviews (PRISMA-ScR): checklist and explanation. Ann Intern Med 2018;169:7:467–73 doi: 10.7326/M18-0850 [first Published Online: 4 September 2018].

23. Jagosh J, Pluye P, Macaulay AC, et al. Assessing the outcomes of participatory research: protocol for identifying, selecting, appraising and synthesizing the literature for realist review. Implement Sci 2011;6:24 doi: 10.1186/1748-5908-6-24 [first published Online: 20 March 2011].

