## Supplementary File - Web of Science - Search Stratergy for "Taking a participatory research approach within workplace health promotion research to improve physical activity levels in office-based workers: a scoping review protocol"

**Co-authors**

Brian P Carson (ORCID iD: 0000-0001-8350-1481)

Department of Physical Education and Sport Science, Physical Activity for Health Research Cluster, Health Research Institute, University of Limerick, Limerick, Ireland

Alan Donnelly (ORCID iD: 0000-0002-6874-0991)

Department of Physical Education and Sport Science, Physical Activity for Health Research Cluster, Health Research Institute, University of Limerick, Limerick, Ireland

Jon Salsberg (ORCID iD: 0000-0003-2010-3691)

Public and Patient Involvement Research Unit, School of Medicine and Health Research Institute, University of Limerick, Limerick, Ireland

**Web of Science – Search Strategy**

ALL=(Health promotion OR Promotion, Health OR Promotions, Health OR Promotion of Health OR Health Promotions OR Promotional Items OR Item, Promotional OR Items, Promotional OR Promotional Item OR Wellness Programs OR Program, Wellness OR Programs, Wellness OR Wellness Program OR Health Campaigns OR Campaign, Health OR Campaigns, Health OR Health Campaign)

AND ALL=(Participatory research OR Community Based Participatory Research OR Participatory Research, Community-Based OR Consumer-Driven Community-Based Research OR Community-Based Research, Consumer-Driven OR Community-Based Researchs, Consumer-Driven OR Consumer Driven Community Based Research OR Consumer-Driven Community-Based Researchs OR Research, Consumer-Driven Community-Based OR Researchs, Consumer-Driven Community-Based)

AND TS=(Employees OR Personnel OR Workers OR Group, Occupational OR Groups, Occupational OR Occupational OR Group OR Employee OR Worker)

AND TS=(Workplace OR Workplaces OR Work Location OR Location, Work OR Locations, Work OR Work Locations OR Work-Site OR Work Site OR Work-Sites OR Work Place OR Place, Work OR Places, Work OR Work Places OR Job Site OR Job Sites OR Site, Job OR Sites, Job OR Worksite OR Worksites)

AND TS=(Walk* OR Step* OR Run* OR Stand* OR Jog* OR Exercises OR Physical Activity OR Activities, Physical OR Activity, Physical OR Physical Activities OR Exercise, Physical OR Exercises, Physical OR Physical Exercise OR Physical Exercises OR Acute Exercise OR Acute Exercises OR Exercise, Acute OR Exercises, Acute OR Exercise, Isometric OR Exercises, Isometric OR Isometric Exercises OR Isometric Exercise OR Exercise, Aerobic OR Aerobic Exercise OR Aerobic Exercises OR Exercises, Aerobic OR Exercise Training OR Exercise Trainings OR Training, Exercise OR Trainings, Exercise)

ALL = All Fields (searches all of the searchable fields using one query. This allows you to easily find your search terms in any field).

TS = Topic (searches title, abstract, author keywords, and keywords plus).
